# University patenting and licensing practices in the United Kingdom during the COVID-19 pandemic

**DOI:** 10.1101/2021.09.20.21263777

**Authors:** Sarai Keestra, Florence Rodgers, Rhiannon Osborne, Sabrina Wimmer

## Abstract

Universities play a vital role in biomedical innovation during the COVID-19 pandemic, and decisions made during technology transfer may affect affordability, accessibility, and availability of health technologies downstream. We investigated the measures the top 35 UK universities receiving most Medical Research Council funding have taken in technology transfer to ensure global equitable access to health technologies. We sent Freedom Of Information (FOI) requests and analysed universities’ websites, to (i.) assess institutional strategies on the patenting and licensing of COVID-19-related health technologies, (ii.) identify all COVID-19-related health technologies licensed or patented, and (iii.) record whether universities engaged with the Open-COVID pledge, COVID-19 Technology Access Pool (C-TAP), or Association of University Technology Managers (AUTM) COVID-19 licensing guidelines. Except for the Universities of Oxford and Edinburgh, UK universities have not updated their institutional strategies during the pandemic. Nine universities licensed 22 COVID-19 health technologies. Imperial College London disclosed 10 patents relevant to COVID-19. No UK universities participate in the Open-COVID Pledge or C-TAP, but discussions are ongoing. The University of Bristol signed up to the AUTM guidelines. Despite several important COVID-19 health technologies being developed by UK universities, our findings suggest minimal engagement with measures that may promote equitable access downstream. We suggest that universities review their technology transfer policies and implement global equitable access strategies for COVID-19 health technologies. We furthermore propose that public and charitable funders can play a larger role in encouraging universities to adopt such practices, by making access and transparency clauses a mandatory condition for receiving public funds for research.

## Introduction

Health equity has previously been defined as “*the absence of systematic disparities in health (or in the major social determinants of health) between social groups who have different levels of underlying social advantage/disadvantage—that is, different positions in a social hierarchy*” (Braveman 2003). Against the backdrop of global vaccine inequalities, in which >60% of the population in high-income countries is vaccinated, compared to <3% in low-income countries, it is essential to re-imagine how health innovation during the pandemic can deliver health equity and public value for all members of the global society, despite evident conflicts between incentivising innovation through exclusive intellectual property (IP) and global health priorities (Keestra 2021; MSF 2021; Thambisetty et al. 2021; UCL-IIPP 2018). In most instances, by transferring the exclusive rights to use intellectual property (IP) around a health innovation to the private sector in the form of an exclusive license, the university relinquishes its influence over the subsequent development and marketization, and thereby the ability to ensure affordability and accessibility downstream (Chokshi 2006; Keestra 2021). Given the urgency of the COVID-19 pandemic, which can only be resolved by equitable access to health technologies (including vaccines, testing kits, ventilators and therapeutics) globally (McMahon 2021), we aimed to identify through Freedom Of Information (FOI) requests the measures universities have taken during the pandemic to protect the human right to health and ensure global equitable access to COVID-19 health technologies developed with public funding. Such measures may include the adoption of new policies or mechanisms aimed at expediting access to COVID-19 IP through implementing specific conditions in university technology transfer to the private sector.

In the UK, where we conducted this study, universities have been at the forefront of health innovation to address COVID-19, as exemplified by the Oxford-AstraZeneca vaccine which originated from the University of Oxford (Cross et al. 2021), with several COVID-19 vaccines under development at the University of Nottingham, Nottingham Trent University (NTU 2020; Zagnat 2021), Imperial College London (Scheuber 2020; UKRI 2021), and the University of Cambridge (University of Cambridge 2021). To support this innovation drive, universities have received significant public funding for COVID-19 related research and development (R&D) to address the SARS-CoV-2 pandemic from the UK government, charities, and public private partnerships, such as the Coalition for Epidemic Preparedness Initiative (CEPI) (Cross et al. 2021; UAEM 2021; UK Research & Innovation 2020). However, significant concerns exist remain regarding global equitable access to COVID-19 health technologies that have been developed at UK universities using significant public financial support, as due to the lack of access conditions attached to such funding, there is currently no national or international measures in place to ensure that publicly funded health innovation delivers public value in the context of global health equity (Cross et al. 2021; Keestra 2021; McDonagh 2021; Pepperrell et al. 2021; UCL-IIPP 2018; Wimmer and Keestra 2020). It has previously been argued that approaches that universities take in managing the IP of health technologies have implications for the accessibility, affordability, and availability for patients globally, and that in the context of global health equity, universities have both the opportunity and the responsibility to ensure that the fruits of publicly funded research meets global public health needs with particular attention towards preventing access barriers in low-and middle income countries (LMICs) (Gotham et al. 2016; Keestra 2021; ‘t Hoen 2003). By allowing for the commercialisation of collective knowledge generated with public investment in the form of exclusive licenses, universities may contribute to the violation the social and economic rights of those unable to access novel biomedical inventions developed at their research institutions (Keestra 2021). However, there are different mechanisms that universities can employ in technology transfer to promote equitable access downstream. For example, the use of non-exclusive licenses, which lets several parties exploit IP, allows for generic competition, which has shown to reduce prices of drugs (Crager, Guillen, and Price 2009; Wiggins and Maness 2004). A notable example is the price decrease in HIV drugs after generic competition was allowed, which contributed to improved global access (MSF Access Campaign 2013). Similarly, by promoting non-exclusive licensing of IP emerging from their COVID-19 research during the pandemic, universities can create an environment in which new health innovations funded by the global public can be optimally disseminated throughout different income contexts and generic competition can lead to affordable pricing. At the same time, it is important that universities, when licensing biologics such as vaccines, realise that not only patents can become an access barrier downstream, but that special measures in technology transfer are needed to ensure access to materials, knowhow, and other proprietary trade secrets (Crager et al. 2009).

During the COVID-19 pandemic a number of mechanisms have emerged for universities to promote global equitable access to COVID-19 health technologies specifically. These mechanisms share a basis in the idea that non-exclusive licensing may enhance access to IP downstream. One such example is the Open-COVID pledge, a Creative Commons led project that calls on organisations to publish a standardised, irrevocable and legally enforceable pledge promising the public free use of their IP in the fight against COVID-19 for the duration of the pandemic as defined by the World Health Organization (WHO) (Open Covid Pledge 2020). This pledge would enable anyone with the capability of doing so to manufacture and distribute such a technology without the threat of litigation and without paying for a license, therefore removing a financial barrier and enabling wider access to such technologies. The limitations of such pledges include the designated time frame after which IP rights are no longer free for all to use, and the subsequent loss of capital for organisations that have set up manufacture of these technologies and are no longer able to do so (Contreras et al. 2020). A second mechanism, the WHO Covid Technology Access Pool (C-TAP) is a voluntary patent pool for sharing IP, know-how, and data related to COVID-19 health technologies with generic manufacturers (McMahon 2021; WHO 2020). The C-TAP builds upon the existing model of the Medicines Patent Pool (Medicines Patent Pool 2021), which negotiates voluntary licenses to promote generic manufacturing and was originally founded to increase access to HIV antivirals (Burrone et al. 2019). The C-TAP can promote access by making it easier for generic manufacturers to secure all licenses needed to produce a technology that may have its IP spread across multiple ‘owners’ if they have all been secured by the pool (McMahon 2021; Thambisetty et al. 2021). However, both the Open Covid Pledge and the C-TAP require industry engagement that has not yet been seen to the extent required (Thambisetty et al. 2021). Universities have the ability to contribute IP directly to these mechanisms and, it could be argued, this would be a suitable public repayment of the money they receive to conduct research that results in licensable technologies (Cross et al. 2021). Additionally, the US-based Association of University Technology Managers (AUTM) have published COVID-19 licensing guidelines aimed at universities specifically that recommend “*time limited, non-exclusive, royalty free licenses”*, which universities can sign up to online (AUTM 2020). Finally, universities also have the possibility to develop their own specific policies or institutional strategies regarding the licensing and technology transfer of COVID-19 health technologies to promote access downstream. Here we aim to give an overview of UK universities’ engagement with different mechanisms available to expedite access to COVID-19 IP, and discuss the implications for global equitable access.

## Methodology

Freedom of information (FOI) requests were filed to 35 UK universities in the period between 24^th^ and 27^th^ October 2020. As we were particularly interested in investigating the technology transfer practices of universities receiving significant amounts of public funding, higher education institutions that received more than 1 million GBP in research grants from the Medical Research Council in 2017-2018 were included, this is the latest year that such funding information is available for (MRC 2019, 2021). In the FOI, universities were asked to disclose information regarding (i.) a list of all COVID-19-related health technologies that had been licensed by the university, including whether the license was exclusive or non-exclusive and in what countries, as well as a list of all COVID-19-related health technologies that had been patented by the university, (ii.) policy changes or institutional strategy on the patenting and licensing of COVID-19-related health technologies, (iii.) whether the university had plans to sign up to the Open-COVID pledge, and/or had been considering licensing particular COVID-19-related health technologies to the C-TAP initiative or the MPP. We additionally searched for policy changes on the universities’ websites and looked at the signatories list of the AUTM COVID-19 guidelines. The full text of the FOI request and university responses are included as supplementary files and publicly available on the digital platform whatdotheyknow.org (S1 & S2).

## Results

Of the 35 universities contacted, 27 universities replied within the legal time frame of 20 working days as stipulated within the Freedom of Information Act (2008), another 8 universities responded with a delay of more than a working week (Newcastle University, University College London, University of Birmingham, University of Cambridge, University of Dundee, University of Edinburgh, University of Sheffield and University of Southampton) (Supplementary 2). We excluded three universities that were not able to disclose licenses and patents related to COVID-19 relevant health technologies in the format (as an excel sheet) as requested for this part of the analysis (Newcastle University, Swansea University, and University of Leeds). The University of Oxford only partially replied to this question. Additionally, the University of Oxford failed to respond to the question about engaging with mechanisms and/or pledges to expedite access to COVID-19 and was therefore excluded from the final part of the analysis.

### Patent and licenses for COVID-19 health technologies

Of the 32 universities that responded to this part of our inquiry, 28% [9/32] reported that they had licensed COVID-19 related health technologies (Imperial College London, University College London, University of Aberdeen, University of Birmingham, University of Bristol, University of Cambridge, University of Oxford, University of Southampton, and the University of Sussex). Together these universities have licensed 22 different health technologies since the start of the pandemic until late October/early November 2020, listed in Table 1. They include 3 vaccines, 6 ventilator related technologies, and 7 licenses related to diagnostics for SARS-CoV2. Of the individual licenses 50% [11/22] were filed by the University of Oxford, which refused to disclose whether these were exclusive or non-exclusive licenses, citing commercial interests as an exemption permitted under FOI law. Of the licenses filed by the other universities 54.5% [6/11] were non-exclusive licenses, whereas 45.5% [5/11] were exclusive.

**Table 1:** Licenses for COVID-19 technologies as disclosed by 32 UK universities autumn 2020.

| University | Technology | Country of Licence | Number of Licences | Exclusive, Non-Exclusive |
| --- | --- | --- | --- | --- |
| Imperial College London | Vaccine | World Wide (WW) | 2 | Exclusive |
| Imperial College London | Ventilator design | World Wide (WW) | 341 | Non-exclusive |
| University College London (UCL) | UCL-Ventura breathing aid (CPAP) – Design and manufacturing package | 122 countries (see Supplementary for further detail) | 1978 | Non-exclusive |
| University of Aberdeen | ATMO-Vent (Atmospheric Mixture Optimisation Ventilator) | Rwanda | 1 | Non-exclusive |
| University of Birmingham | Anti-IgG/IgA/IgM SARS-CoV2 Spike Protein assay | World Wide (WW) | 1 | Exclusive |
| University of Bristol | Potential SARS-CoV2 target, screening assay and therapeutic strategy | World Wide (WW) | 1 | Exclusive |
| University of Cambridge | Biological vaccine | World Wide (WW) | 1 | Exclusive |
| University of Oxford | Novel coronavirus vaccine | Did not disclose citing section 43(2) of the Freedom Of Information Act (FOIA), which states that information is exempt where its disclosure would, or would be likely to, prejudice the commercial interests of any person. The University of Oxford says in their FOI response: "*It is our view that disclosure would be likely to prejudice the commercial interests of Oxford University Innovation (OUI) and/or the University, because the information would weaken the bargaining position of the OUI/University in negotiating similar agreements with potential licensees in future.*" The complete reasoning for non-disclosure is attached in the Supplementary. |  |  |
| University of Oxford | Coronavirus testing primer design |  |  |  |
| University of Oxford | Rapid test method for coronavirus |  |  |  |
| University of Oxford | Ventilator electronics design |  |  |  |
| University of Oxford | Mechanical ventilator |  |  |  |
| University of Oxford | Ventilator software control system |  |  |  |
| University of Oxford | ChAdOx2 - simian adenovirus vector |  |  |  |
| University of Oxford | ChAdOx1 A new adenoviral vector |  |  |  |
| University of Oxford | Adenovirus long promoter |  |  |  |
| University of Oxford | Rapid test method for Coronavirus |  |  |  |
| University of Oxford | Coronavirus testing primer design |  |  |  |
| University of Southampton | Interferon Beta technology | World Wide (WW) | 1 | Exclusive |
| University of Southampton | A Personal Respirator for Healthcare Professionals Treating COVID-19 (PeRSo) | World Wide (WW) | NS | Non-exclusive |
| University of Sussex | Alcohol-based hand rub | UK and Zambia | NS | Non-exclusive |
| University of Sussex | Diagnostic testing systems | UK | NS | Non-exclusive |

Only one university (Imperial College London) provided a list of patents relevant for COVID-19, which included a ventilator, a hand wash device, an antibody assay, a vaccine, and six method and apparatus patents, filed in the UK or the Patent Cooperation Treaty (PCT). They noted however that “*unlike other Patent Offices, such as the United States Patent and Trademark Office, the UKIPO has yet to launch a formal scheme offering applicants prioritized examination of COVID-19 cases*.” Other universities said that no patents on COVID-19 health technologies had been granted to them as of late October/early November 2020.

### Policy statements on technology transfer during the COVID-19 pandemic

Of the 35 Universities included in this analysis, one university, the University of Oxford, has released an institutional default approach regarding patenting and licensing of COVID-19 related health technologies in a statement titled “Expedited Access for COVID-19 IP” which was released in Spring 2020 (Oxford University Innovation 2020). The other 34 universities (97%) did not have a COVID-19 specific patenting and licensing strategy. One university, the University of Edinburgh, updated their internal strategy on patenting and licensing in a university-wide revised essential medicines policy in September 2020 (The University of Edinburgh 2020, 2021). This policy is not limited to COVID-19 related health technologies alone but covers all health technologies developed through research at the university.

### Use of the Open COVID Pledge, C-TAP, and the AUTM guidelines for COVID-19 technologies as mechanisms to expedite access

None of the 34 universities included in this part of the analysis (which excludes the University of Oxford, who failed to respond to this part of the FOI) have adopted the Open COVID Pledge or used C-TAP as a mechanism to expedite access to COVID-19 related health technologies. However, three universities (Birckbeck, King’s College London, and Swansea University) said autumn 2020 that review of the Open COVID Pledge is ongoing. Additionally, three universities (Queen’s University Belfast, University of Birmingham, University of Bristol) mentioned that although they currently do not have COVID-19 related health technologies to license to the C-TAP initiative, they would consider licensing to the C-TAP on a case-by-case basis should they develop these in the future. Finally, one university (University of Cambridge) disclosed that they have already had discussions directly with the WHO C-TAP/MPP initiative, but did not consider the mechanism appropriate for all health technologies and would not usually seek patents outside of high-income countries anyway. The University of Bristol is the only UK university that signed up to the AUTM guidelines for COVID-19 health technologies (AUTM 2020).

## Discussion

We conducted an assessment of 35 UK universities receiving the most MRC funding in the year 2017-2018, as a proxy of significant engagement with public funds, looking at their patenting and licensing practices regarding COVID-19 health technologies in the context of promoting global equitable access downstream. Of the licenses for which exclusive status was known, 54.5% were non-exclusive, which is higher than the 30% recorded in a similar sample of UK universities before the COVID-19 pandemic started (Gotham et al. 2016). Prior to the pandemic, seven universities in the UK had implemented an essential medicines policy or committed to the principles of socially responsible licensing, including University College London, Imperial College London and the Universities of Oxford, Dundee, Bristol, and Edinburgh. These policies included measures such as refraining from prosecuting patent applications in developing countries and using non-exclusive licenses in ways that promote access (Gotham et al. 2016). In this study we identified six non-exclusive licenses for COVID-19 health technologies developed by UK universities, three of which concerned ventilator designs. Health technologies that were licensed in LMICs specifically were all non-exclusive. In contrast, assays, vaccines, and potential therapeutic targets were mostly licensed exclusively, which includes the COVID-19 vaccines developed by the University of Cambridge, Imperial College London and the University of Oxford. This is particularly concerning as in 2021, inequalities in vaccine access across income contexts due to limited supply and affordability issues remains a painfully real display of the inequities in our biomedical innovation system (Keestra 2021; MSF 2021). Despite initial commitments by the University of Oxford to non-exclusive licensing (Oxford University Innovation 2020), the University later entered into an exclusive licensing deal with the British-Swedish pharmaceutical company AstraZeneca to further the development of the ChAdOx1 nCoV-2 vaccine (U.S. House of Representatives 2020). Still, the Oxford-AstraZeneca is one of the most affordable COVID-19 vaccines, suitable for deployment in LMICs, and in its technology transfer deal University of Oxford has committed AstraZeneca to sell the vaccine at cost-price in perpetuity in the lowest income countries (le Moel 2020; Safi 2021). Although it is important to acknowledge the steps University of Oxford thereby has taken to promote accessibility downstream, vaccine shortages, inequitable global access, and the vaccine price post-pandemic in LMICs remains a concern (Keestra 2021; Safi 2021). Imperial College London chose another mode of technology transfer by exclusively licensing its COVID-19 vaccine IP to a newly established social enterprise, “VacEquity Global Health”, which has committed to waiving all royalties in the UK and LMICs to promote equitable access in resource limited settings (Scheuber 2020). As the vaccine is still undergoing development the implementation of these commitments in practice remains to be seen. It is unknown till date through which technology transfer modality the Universities of Cambridge and Nottingham will further commercialise their vaccines for COVID-19.

We found that the majority of universities had not changed their technology transfer strategies in response to the pandemic. The lack of adoption of new technology transfer policies may be because universities felt that their existing patenting and licensing practices were adequate, and/or gave sufficient scope to make unique decisions for different health technologies depending on the circumstance. Analysis of how existing policies correlated with different decisions on COVID-19-related health technology patenting and licensing strategies would therefore be of interest for future research. There were two notable exceptions of universities that had adopted novel technology transfer practices at the beginning of the COVID-19 pandemic. In Spring 2020, Oxford University’s technology transfer office, Oxford University Innovation (OUI) released a statement noting that “*the COVID-19 pandemic demands an urgent and unprecedented response*” and that “*university research and expertise is critical to this effort*” (Oxford University Innovation 2020). OUI subsequently specified that their default strategy for the technology transfer for COVID-19 related IP would be to “*offer non-exclusive, royalty-free licences to support free of charge, at-cost or cost + limited margin supply as appropriate, and only for the duration of the pandemic, as defined by the WHO”*. Although commendable in principle, it remains unclear whether these commitments were adhered to in practice, due to a lack of transparency in the terms of the AstraZeneca vaccine deal. Furthermore, this statement included no access provisions for the technology transfer of biologics such as vaccines, where additional measures need to be taken to ensure access to materials and knowhow needed to manufacture such health technologies (Crager et al. 2009). Although the early R&D of the ChAdOx vector that is used in this vaccine technology was largely developed using public funds (Cross et al. 2021), the contract we have received through FOI is heavily redacted, particularly its access clauses (Safi 2021). This lack of transparency and secrecy around the commercialization of publicly-funded health research is an ongoing problem throughout the biomedical innovation system, and is reprehensible as it hinders public accountability from recipients of such funding (Keestra 2021; UCL-IIPP 2018). To ensure that public financing of health research delivers public value, increased transparency from universities and their commercial partners is essential. This is a key feature of the University of Edinburgh updated its Essential Medicines Position Statement applicable to the IP of all health technologies originating at the university, and is not limited to COVID-19 health technologies alone (The University of Edinburgh 2020). In this Position Statement they commit to *“making information public concerning negotiations regarding technology transfer with a third party (…) publishing technology transfer agreements in full for all health technologies (…) [and the] establishment of a committee that monitors and promotes the adherence to the goals and provisions committed to by the University of Edinburgh in this policy*.*”* Other universities similarly have the opportunity to implement radical transparency in technology transfer deals, and research funders could make this a condition of receiving public funds.

We found that none of the UK universities included in our cohort had adopted existing other mechanisms to expedite equitable access to COVID-19 health technologies, such as the Open Covid Pledge, C-TAP, or the AUTM guidelines. We see two main reasons for this, firstly, that some universities did not find these standardised measures appropriate to cover all of their health technology transfer modalities during the pandemic, and secondly, that these mechanisms are completely voluntary and rely only on the good nature of the patent-holders to commit and uphold the standards of these voluntary licensing mechanisms (McMahon 2021). Signatories to the Open-COVID pledge are mostly technology, software, or social media companies, such as IBM, Amazon, Microsoft and Facebook (Open Covid Pledge 2021), and as of yet, no university in the UK has committed to or endorsed the pledge. This suggests that perhaps this mechanism is less suitable for universities and their existing approaches regarding technology transfer. This might be due to concerns by a universal IP pledge may inadvertently affect licensing agreements for COVID-19 related IP that were signed prior to the COVID-19 pandemic. In contrast, the C-TAP, which was launched by the WHO’s Access to COVID-19 Tools (ACT) Accelerator (ACT-A), has been specifically designed to accommodate for the voluntary sharing of COVID-19 health technologies’ IP and knowhow of. It is intended to particularly ease technology transfer to generic and biosimilar manufacturers in the Global South to increase manufacturing and global equitable access (Correa 2021; WHO 2021). However, no COVID-19 health technologies had been licensed to C-TAP at the time when we conducted our study in late October 2020. We suspect that perhaps because health innovations originating from universities are often still at an early research stage, they may sometimes not be ready to be added to the C-TAP without further clinical trials in humans. There is currently little opportunity for universities to carry such research out themselves without a private sector partner. However, UK universities and other research institutions should consider adding clauses to technology transfer contracts with the private sector demanding the sharing of IP with the C-TAP downstream as a mechanism to promote access to publicly funded COVID-19 health technologies. Indeed, the Medicines Patent Pool (MPP), which inspired the creation of C-TAP was granted its first voluntary license by a public research institution, which has set the historical precedent that universities can play an important role in encouraging the use of patent pooling mechanisms. In 2010, the US National Institutes of Health and co-patent owner the University of Illinois at Chicago granted the pool a license for a HIV-drug (Hillary Chen 2010). Further, in 2017, Johns Hopkins licensed the first tuberculosis drug to the MPP (Medicines Patent Pool 2017), which helped to get political will behind the mechanism. UK universities have the opportunity to play a similarly important role in the current pandemic. Finally, most current signatories of the AUTM guidelines are universities based in the United States, with the notable exception of the University of Bristol (AUTM 2020). This is unsurprising as AUTM is a US-based organisation, which only receives limited engagement from UK universities. The pandemic has highlighted the need of an official and transparent UK-specific equivalent of AUTM, which could coordinate the sharing of best practices around non-exclusive licensing and specific access clauses for inclusion in technology transfer deals. There is currently no oversight or coordinating body that ensures that publicly funded research conducted by universities reaches the global public on an equitable basis, although there is moral imperative to do so (Keestra 2021). Instead, the current biomedical innovation system relies on the good will of its actors to commit voluntarily to the mechanisms outlined here (McMahon 2021), whereas it might be more effective for public and charitable funders to make it mandatory for universities to adhere to certain access and transparency conditions when developing novel health innovations that arise from public financing of research.

Our study shows that utilising the Freedom Of Information Act is a useful methodology to gain insight into the role of universities in the R&D landscape that has emerged during the COVID-19 pandemic. However, engaging with FOIs as a research method has limitations, as public institutions are allowed to refuse requests under section 43(2) of The Act if “*its disclosure under this Act would, or would be likely to, prejudice the commercial interests of any person (including the public authority holding it)*” (Freedom of Information Act 2000). This exemption based on commercial interests is sometimes employed by public research institutions to justify non-disclosure of information regarding the commercialisation of health technologies, as it is deemed commercially sensitive. In the case of this particular study, the University of Oxford stated that disclosure of exclusivity status of licenses and patents under application would weaken the bargaining position of the OUI/University in the future and argued that this is also against public interest (Supplementary 2). We remain concerned that this lack of transparency by public research institutions, such as universities, which receive large amounts of public funding, hinders accountability whilst engagement of public and charitable entities with the private sector should arguably be subjected to public scrutiny through mechanisms such as the Freedom Of Information Act (2000).

## Limitations

Our analysis did not include discussion of whether licenses, both exclusive or non-exclusive, contained any further access maximising conditions or clauses such as pricing conditions, sub-licensing requirements, or step-in rights. Furthermore, our study did not explore which types of technology transfer are suitable for different health technologies, and what the outcomes of different types of technology transfer are. Biologics such as COVID-19 vaccines for example, will likely require different considerations in technology transfer to ventilator designs or small molecule drugs. A further challenge encountered in our study methodology was the lack of transparency regarding technology transfer, as some universities did not disclose information in their response to our FOI request due to commercial interests.

## Conclusion

Considering the unique position of universities in the biomedical innovation ecosystem, situated between upstream publicly funded R&D and often downstream private sector marketisation, universities have both the opportunity and responsibility to determine the conditions of technology transfer (Keestra 2021). This has especially been important in the response to the COVID-19 pandemic, in which public research institutions and public funding played a prominent role. Universities defy the essential principles they were founded upon when contributing to the global tragedy of preventable deaths during the COVID-19 pandemic when they fail to consider implications for downstream access in their technology transfer practices (Keestra 2021). We therefore encourage universities to review their patenting and technology transfer policies and practices for COVID-19 technologies and commit to promoting global equitable access, through mechanisms such as the Open-COVID pledge, C-TAP, and the AUTM COVID-19 licensing guidelines, or other ways to promote non-exclusive licensing and access protections, such as outlined in the University of Edinburgh’s Essential Medicines Position Statement (The University of Edinburgh 2020). Universities can also further develop institutional COVID-19 specific policies regarding technology transfer as the University of Oxford did, and commit to support royalty-free licenses and at-cost supply of key health technologies during the COVID-19 pandemic. Beyond the pandemic, we additionally recommend that universities commit to increased transparency in technology transfer, and that public and charitable funders can play a larger role in encouraging universities to adopt such practices, by making access and transparency clauses a mandatory condition for receiving public funds for research. It is important that we continue to develop a health innovation landscape that is open to the adoption of novel strategies that encourage global equitable access to health technologies for all, everywhere, during this pandemic and beyond.

## Supporting information

whatdotheyknow.org (S1 & S2).

whatdotheyknow.org (S1 & S2).

## Data Availability

Universities responses are available in Supplementary 2. The FOIs are also publicly accessible online via WhatDoTheyKnow.com. For further information please contact the corresponding author.

https://www.whatdotheyknow.com/user/sarai_keestra

https://www.whatdotheyknow.com/user/florence_rodgers?page=2

