## Supplementary material for "University patenting and licensing practices in the United Kingdom during the COVID-19 pandemic": whatdotheyknow.org (S1 & S2).

Dear UNIVERSITY,

For the purpose of clarity, please adhere to the following definitions when answering these questions:

Low and middle income country: A country that is defined as ‘low’, ‘lower-middle’ and ‘upper-middle’ income by the World Bank for the year 2018. Income status defined by GNI per capita.
Source: World bank, available online [[https://tinyurl.com/y5dwc2qv]](https://tinyurl.com/y5dwc2qv%5D)

A patent grants an inventor the exclusive rights to his or her invention. A patent holder can stop other people from selling, manufacturing, producing, or using the invention for a certain period of time.
Source: Upcouncil, available online [<https://www.upcounsel.com/what-is-a-pate>...

Licence: Permission for the licensee to use the intellectual property rights
Source: TaylorWessing, available online [<https://www.taylorwessing.com/download/a>...

Exclusive Licence: A licence that allows only the named licensee to exploit the relevant IP rights, the licensor is also excluded from exploiting these IP rights.
Non-Exclusive Licence: A licence that grants any number of licensees to exploit the IP.
Source: TaylorWessing, available online [<https://www.taylorwessing.com/synapse/co>...

Health Technology: ‘Health technology as defined by the WHO: “A health technology is the application of organized knowledge and skills in the form of devices, medicines, vaccines, procedures and systems developed to solve a health problem and improve quality of lives.’
Source: WHO, available online [<https://www.who.int/health-technology-as>...

START FOI request:
Q1. Can you release your internal strategy regarding licensing and patenting of COVID-19 related health technologies?

Q2. Does the institution have a formal policy on licensing and patenting of COVID-19 related technologies? If yes, please provide a link or PDF to the relevant policy document.

Q3. Has the university considered updating their strategy regarding the licensing and patenting of health technologies since the start of the pandemic?

Q4. Please provide an Excel spreadsheet with headings as displayed below with all the COVID-19 related health technologies LICENSED by the university AND specify which countries they were licensed in AND specify whether they are exclusive or non-exclusive licenses.
Health Technology Licensed | Country of Licensing | Type of License (Exclusive or Non-Exclusive)

Q5. Please provide a list of patents granted to your organization for any COVID-19 related health technologies listed by patent family, indicating countries/regions in which the patent has been granted.

Q6. Does the institution have any plans on signing up to the Open-COVID pledge (<https://opencovidpledge.org/>) or any similar initiative that seeks to minimise intellectual property rights barriers during the COVID-19 pandemic? If yes, please specify.

Q7. Is the institution considering the licensing of COVID-19 related health technologies to the Covid Technology Access Pool (C-TAP) or the Medicines Patent Pool (MPP) during the pandemic? If yes, please specify.

END FOI request.
If you have any questions please do not hesitate to contact me by email.

Yours faithfully,

SENDER NAME
